# Reduced Vestibular Function is Associated with Cortical Surface Shape Changes in the Frontal Cortex

**DOI:** 10.1101/2024.11.22.24317807

**Authors:** Dominic Padova, J. Tilak Ratnanather, Andreia V. Faria, Yuri Agrawal

**Author notes:** Corresponding author: Dominic Padova.

## Abstract

Aging-associated decline in peripheral vestibular function is linked to deficits in executive ability, self-motion perception, and motor planning and execution. While these behaviors are known to rely on the sensorimotor and frontal cortices, the precise pathways involving the frontal and sensorimotor cortices in these vestibular-associated behaviors are unknown. To fill this knowledge gap, this cross-sectional study investigates the relationship between age-related variation in vestibular function and surface shape alterations of the frontal and sensorimotor cortices, considering age, intracranial volume, and sex. Data from 117 participants aged 60+ from the Baltimore Longitudinal Study of Aging, who underwent endorgan-specific vestibular tests (cVEMP for the saccule, oVEMP for the utricle, and vHIT for the horizontal canal) and T1-weighted MRI scans on the same visit, were analyzed. We examined ten brain structures in the putative “vestibular cortex”: the middle-superior part of the prefrontal cortex (SFG_PFC), frontal pole (SFG_pole), and posterior pars of the superior frontal gyrus (SFG), the dorsal prefrontal cortex and posterior pars of middle frontal gyrus (MFG_DPFC, MFG), the pars opercularis, pars triangularis, and pars orbitalis of the inferior frontal gyrus, as well as the precentral gyrus and postcentral gyrus (PoCG) of the sensorimotor cortex. For each region of interest (ROI), shape descriptors were estimated as local compressions and expansions of the population average ROI surface using surface LDDMM. Shape descriptors were linearly regressed onto standardized vestibular variables, age, intracranial volume, and sex. Lower utricular function was linked with surface compression in the left MFG and expansion in the bilateral SFG_pole and left SFG. Reduced canal function was associated with surface compression in the right SFG_PFC and SFG_pole and left SFG. Both reduced saccular and utricular function correlated with surface compression in the posterior medial part of the left MFG. Our findings illuminate the complexity of the relationship between vestibular function and the morphology of the frontal and sensorimotor cortices in aging. Improved understanding of these relationships could help in developing interventions to enhance quality of life in aging and populations with cognitive impairment.

## 1 Introduction

The five organs of the peripheral vestibular system, the saccule, the utricle, and the three semi-circular canals, send information about self-motion relative to gravity to a widespread network of multi-sensorimotor brain regions [1, 2, 3, 4, 5]. The vestibular network is involved not only in the maintenance of balance, posture, and stable vision, but also in autonomic, affective, and higher-order behaviors [6]. Additionally, vestibular function has been linked with neurodegenerative diseases that impact these functions, such as dementia [7, 8, 9, 10], multiple sclerosis [11], Huntington’s disease [12], and Parkinson’s disease [13, 14, 15, 16, 17, 18, 19, 20]. Given that vestibular structure and function are known to decline with aging [21, 22, 23, 24, 25, 26, 27], age-related vestibular dysfunction may play a role in balance and cognitive phenotypes in aging and disease. In older adults, age-related vestibular loss is related to deficits in higher-order behaviors, such as attention, visuospatial cognitive ability, executive ability, memory, self-motion perception, and motor planning and execution [28, 29, 30, 31, 32, 33]. For all our expanding knowledge of the relationship between age-related vestibular loss and both higher-order behaviors and neurodegenerative diseases, significant gaps exist in our understanding of the involved neuroanatomical circuits.

The postcentral gyrus, precentral gyrus, and the frontal cortex are vital regions in the vestibular cognitive network [6]. The postcentral and precentral gyri are involved in sensorimotor function, and the prefrontal cortex is involved in executive function. These regions receive and process vestibular and multi-sensorimotor information, including hearing, vision, and proprioception, via thalamo-cortical and cortico-cortical pathways [1, 5, 3]. However, the evidence of the relationships between peripheral vestibular function and the structures of the postcentral gyrus, precentral gyrus, and prefrontal cortex has been inconsistent [34, 35, 36, 37, 38, 39, 40, 41, 42]. Several studies have identified structural alterations in the somatosensory [34, 41, 42], motor [39, 41], and prefrontal cortices [37, 39, 41, 42] with vestibular dysfunction. Furthermore, previous studies of age-related end-organ functions did not examine these multi-sensorimotor regions [43, 44, 45].

To fill these knowledge gaps, we used MRI scans, vestibular, hearing, vision, and proprioception physiologic data from 117 healthy, older adults from the Baltimore Longitudinal Study of Aging to answer two questions:

1. Is age-related vestibular function related to prefrontal and sensorimotor cortex surface morphology in healthy, older adults?
2. Do vestibular-associated morphological alterations in the prefrontal and sensorimotor cortices persist after accounting for multisensory involvement?

This cohort and its measurements were used in previous studies by our group [43, 44] that explored distinct research questions involving a different cognitive network. We hypothesized that higher functioning of the saccule, utricle, and horizontal semi-circular canal is related to surface shape alterations in the regions of interest, even after accounting for multi-sensory function (hearing, vision, and proprioception). This study significantly extends our previous vestibular-only study of prefrontal and sensorimotor volumes, as shape can vary in more complicated, local patterns than does gross volume [42]. This study will improve the understanding of the consequences of aging on the vestibular pathways involved in vestibular cognition. An improved understanding will aid in developing rational strategies to preserve vestibular-mediated behaviors in aging and disease.

## 2. Data and methods

### 2.1 Study sample

The data is a subset of 117 healthy older (aged ≥ 60 years) participants from the Baltimore Longitudinal Study of Aging (BLSA) who had MRI brain scans and vestibular testing in the same visit between 2013 and 2015 [46]. All participants provided written informed consent. The BLSA study protocol (03-AG-0325) was approved by the National Institute of Environmental Health Sciences Institutional Review Board. Hearing loss, visual acuity loss, and proprioceptive loss were measured and included as confounding variables in follow-up hypothesis tests. Hearing loss was measured as the speech-frequency pure tone average of air-conduction thresholds at 0.5, 1, 2, and 4 kHz from the better ear. Visual acuity loss, which refers to how much a pattern must differ in size to be seen, was measured as the angular deviation in logMAR units and ranges from 0.80 to -0.30 logMAR, where lower values indicate better acuity. Proprioceptive loss was measured as the degree of ankle deflection perceptible according to an established BLSA procedure [47]. For analysis, the hearing, vision, and proprioceptive variables were treated as continuous variables and were negated so that increasing values indicate better function.

### 2.2 Vestibular physiologic testing

Vestibular function testing included measurement of saccular function using the cervical vestibular-evoked myogenic potential (cVEMP) test, of utricular function using the ocular VEMP (oVEMP) test, and of horizontal semicircular canal function using the video head-impulse test (vHIT), following established procedures [7, 48, 49, 50].

### 2.2.1 Cervical vestibular-evoked myogenic potential (cVEMP) test

The cVEMP test measures the function of the saccule (and inferior vestibular nerve) [7, 48, 49, 50]. Participants sat on a chair inclined at 30° above the horizontal plane. Trained examiners positioned EMG electrodes bilaterally on the sternocleidomastoid and sternoclavicular junction, with a ground electrode on the manubrium sterni. Participants were instructed to turn their heads to generate at least a 30 *µ*V background response prior to delivering sound stimuli. Bursts of 100 auditory stimuli stimuli of 500 Hz and 125 dB were administered monoaurally through headphones (VIASYS Healthcare, Madison, WI). cVEMPs were recorded as short-latency EMGs of the inhibitory response of the ipsilateral sternocleidomastoid muscle. To calculate corrected cVEMP amplitudes, nuisance background EMG activity collected 10 ms prior to the onset of the auditory stimulus were removed. The higher corrected cVEMP amplitude (unitless) from the left and right sides was used as a continuous measure of saccular function. A difference of 0.5 in corrected cVEMP is considered clinically relevant [48].

### 2.2.2 Ocular vestibular-evoked myogenic potential (oVEMP) test

The oVEMP test measures the function of the utricle (and superior vestibular nerve) [7, 48, 49, 50]. Participants sat on a chair inclined at 30° above the horizontal plane. Trained examiners placed a noninverting electrode ≈3 mm below the eye centered below the pupil, an inverting electrode 2 cm below the noninverting electrode, and a ground electrode on the manubrium sterni. To ensure that symmetric signals are recorded from both eyes, participants were instructed to perform multiple 20° vertical saccades before stimulation. During oVEMP testing, participants were instructed to maintain an upward gaze of 20°. Head taps (vibration stimuli) applied to the midline of the face at the hairline and ≈30% of the distance between the inion and nasion using a reflex hammer (Aesculap model ACO12C, Center Valley, PA). oVEMPs were recorded as short-latency EMGs of the excitation response of the contralateral external oblique muscle of the eye. The higher oVEMP amplitude (*µ*V) from the left and right sides was used as a continuous measure of utricular function. A difference of 5 *µ*V in oVEMP is considered clinically relevant [48].

### 2.2.3 Video head impulse test (vHIT)

The vHIT measures the horizontal vestibular-ocular reflex (VOR) [7, 51, 52] and was performed using the EyeSeeCam system (Interacoustics, Eden Prarie, MN) in the same plane as the right and left horizontal semicircular canals [52, 53, 54]. To position the horizontal canals in the plane of stimulation, trained examiners tilted the participant’s head downward 30° below the horizontal plane and instructed participants to maintain their gaze on a wall target ≈1.5 m away. The examiner delivered rotations of 5-10° (≈150-250° per second) to the participant’s head. The head impulses are performed at least 10 times parallel to the ground toward the right and left, chosen randomly for unpredictability. The EyeSeeCam system quantified eye and head velocity. VOR gain was calculated as the unitless ratio of the eye velocity to the head velocity. A VOR gain equal to 1.0 is normal and indicates equal eye and head velocities. The mean VOR gain from the left and right sides was used as a continuous variable. A difference of 0.1 in VOR gain is considered clinically relevant [7, 48].

### 2.3 Structural MRI acquisition

T1-weighted volumetric MRI scans were acquired in the sagittal plane using a 3T Philips Achieva scanner at the National Institute on Aging Clinical Research Unit. The sequence used was a T1-weighted image (WI) (magnetization prepared rapid acquisition with gradient echo (MPRAGE); repetition time (TR)=6.5 ms, echo time (TE)=3.1 ms, flip angle=8°, image matrix=256×256, 170 slices, voxel area=1.0×1.0 mm, 1.2 mm slice thickness, FOV=256×240 mm, sagittal acquisition). Scans were automatically segmented using MRICloud (https://www.mricloud.org/) with the T1 multi-atlas set “BIOCARD3T_297labels_10atlases_am_hi_erc_M2_252_V1”.

### 2.4 MRI processing pipeline

Our analysis focuses on the ten regions of interest (ROIs) relevant to our hypothesis and shown in Figure 1. These ROIs include the middle-superior part of the prefrontal cortex (SFG_PFC), frontal pole (SFG_pole), and posterior pars of the superior frontal gyrus (SFG), the dorsal prefrontal cortex and posterior pars of middle frontal gyrus (MFG_DPFC, MFG), the pars opercularis, pars triangularis, and pars orbitalis of the inferior frontal gyrus (IFG), as well as the precentral gyrus (PrCG), postcentral gyrus (PoCG) of the sensorimotor cortex. Intracranial volume was comprised of bilateral cerebral volumes, cerebellum, brainstem, and cerebrospinal fluid. We followed a procedure similar to those described in previous studies investigating sub-cortical changes associated with mild cognitive impairment and Alzheimer’s disease [55, 56, 57], Huntington’s disease [58], attention deficit hyperactivity disorder [59], and schizophrenia [60, 61]. Figure 2 depicts an overview of the neuroimaging pipeline.

**Figure 1.**
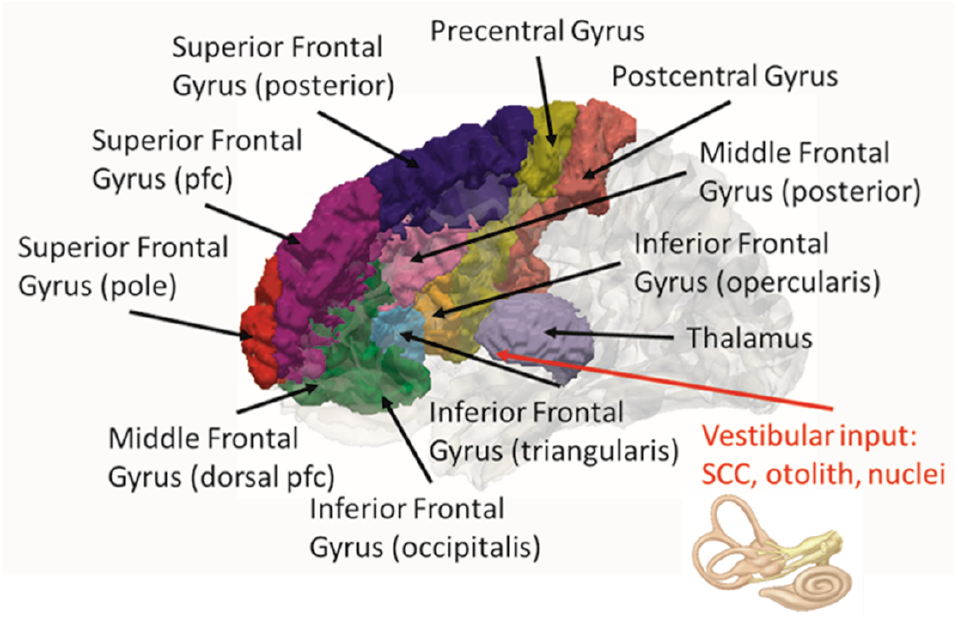
Putative vestibular-thalamocortical and cortico-cortical circuits. Vestibular information from the semicircular canals, otoliths, and vestibular nuclei reaches the precentral and postcentral gyri of the sensorimotor cortex and the frontal gyrus via the thalamo-cortical and cortico-cortical circuits. The red arrow indicates the ventral lateral nucleus of the thalamus which putatively receives vestibular input. CAWorks (www.cis.jhu.edu/software/caworks) was used for visualization. Key: pfc: prefrontal cortex; SCC: semicircular canals.

**Figure 2.**
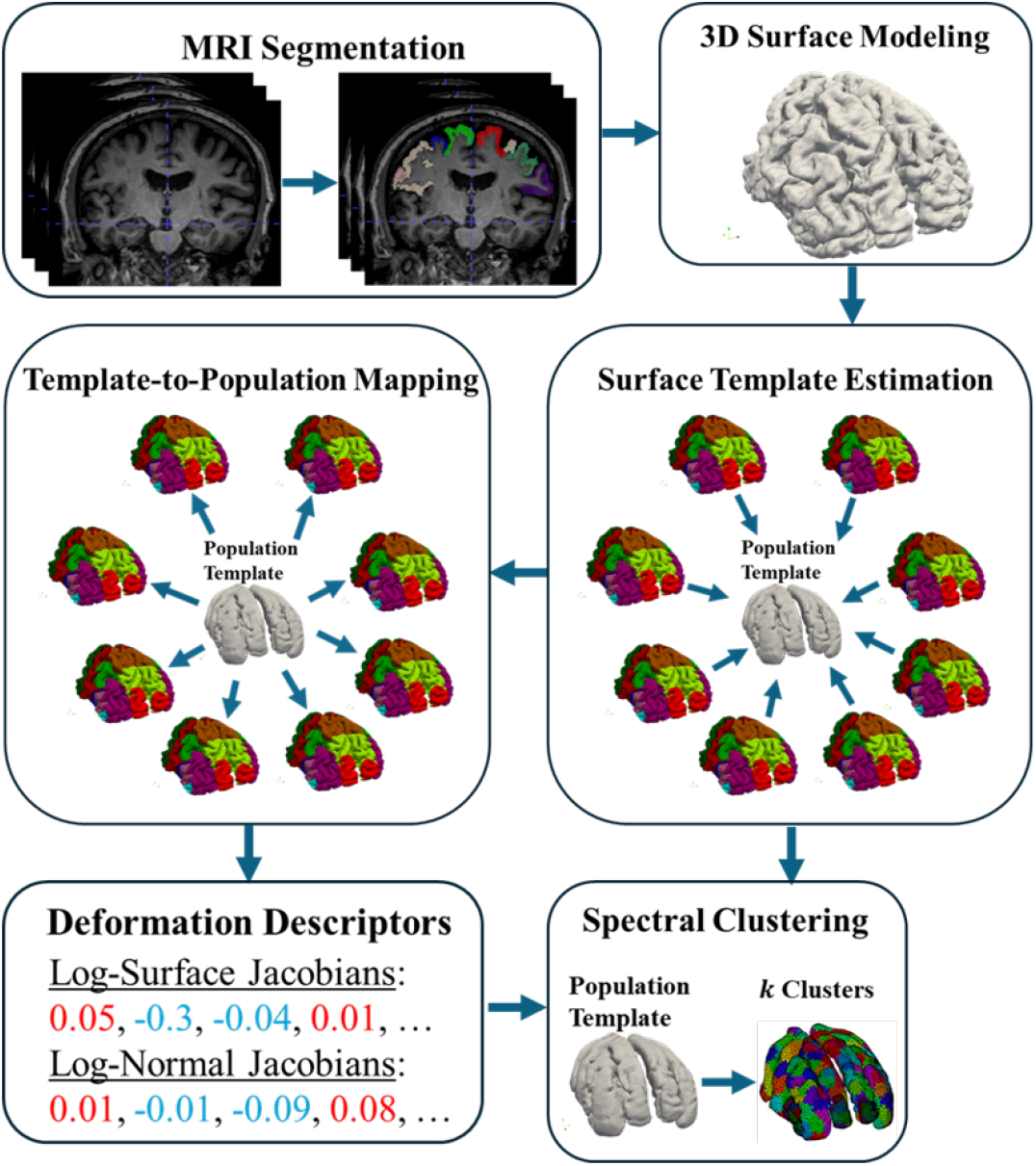
Neuroimaging pipeline. Using T1-weighted MRI scans as inputs, MRICloud automatically outputs a whole-brain parcellation using a study-appropriate multi-atlas and LDDMM. Taking the binary image segmentations of the subset of regions of interest (eight subregions of the frontal cortex, the precentral gyrus, and the postcentral gyrus), 3D surfaces for each structure created using a restricted Delaunay triangulation. Then the collection of surfaces for each structure is uploaded to the MRICloud Shape Analysis pipeline [62, 63] to perform surface template estimation and subsequently template-to-population mapping. The output vertex-wise deformation descriptors (the logarithms of the surface and normal Jacobians) are then reduced to *k* descriptors based on spectral clustering for downstream statistical testing. Quality control was performed at each stage.

### 2.4 Shape analysis

For each 3D segmented ROI, surface meshes with ≈ 800 vertices were generated using a restricted Delaunay triangulation. Using the MRICloud surface template generation pipeline, the collection of ROI surfaces was used to estimate left- and right-side population templates (i.e. the average shape) agnostic to diagnostic criteria by an LDDMM-based surface template estimation procedure after rigid alignment [64]. The MRICloud template-to-population surface mapping pipeline was used to register each participant’s surface to the population template, first rigidly then diffeomorphically using surface LDDMM [64]. Surface shape alterations were measured by the logarithms of surface and normal Jacobian determinants of the diffeomorphic transformation at each vertex. The surface Jacobian is calculated as the ratio between the surface area of the faces attached to a vertex pre- and post-transformation. The normal Jacobian is the ratio between the full Jacobian and the surface Jacobian. Whereas the surface Jacobian refers to change in surface area pre- and post-transformation, the normal Jacobian refers to the change in normal distance pre- and post-transformation. A positive (negative) surface log-Jacobian value denotes an expansion (contraction) of the template around that vertex in the direction tangent to the surface to fit the subject. Similarly, a positive (negative) normal Jacobian value denotes an expansion (contraction) of the template around that vertex in the direction normal to the surface to fit the subject. We analyze the surface and normal log-Jacobians independently. To increase the power of the analyses and to improve computational efficiency, the surfaces were spectrally clustered into *k* ∈ {10, …, 20} clusters of size ≈150-400 *mm*^2^ based on the surface geometry of the template, as described previously [58]. Thus, the *k* shape descriptor variables attached with each subject structure were used as separate outcome variables for hypothesis testing.

### 2.6 Statistical modeling

For participants with missing vestibular data, we carried over data from an adjacent prior or subsequent visit using an external longitudinal dataset comprised of the same participants [65]. Whereas our original dataset had 58, 64, and 91 observations for cVEMP, oVEMP, and VOR, respectively, the imputed dataset had 95, 100, 107 observations for cVEMP, oVEMP, and VOR, respectively. Using this imputed dataset, multiple linear regression adjusted for age, intracranial volume, and sex was used to investigate the relationship between local shape descriptors and vestibular function. The null hypothesis, *H*_0*A*_, in Eq. (1) predicts the (normal, surface) Jacobian *jac*_*i*_, for participant *i, i* = 1, …, *N*. The alternate hypothesis *H*_1_, predicts the (normal, surface) Jacobian *jac*_*i*_ using a vestibular variable, *vest*_*i*_ in Eq. (2), such as best corrected cVEMP, best oVEMP, and mean VOR gain as continuous independent variables,

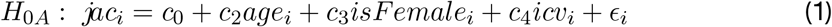

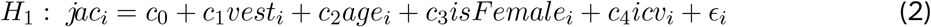

To test whether the addition of the function of hearing, vision, or proprioception either explains away or masked vestibular relationships, we performed three additional bivariate sensory hypothesis tests. The null hypothesis, *H*_0*B*_, in Eq. (3) and the alternative hypothesis, *H*_2_, in Eq. (4) additionally covary for the *sensory* variable, which represents hearing function, vision function, or proprioceptive function,

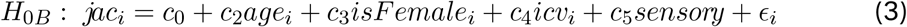

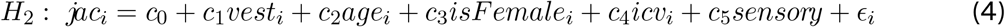

In hypothesis tests{*H*_0*A*_, *H*_0*B*_, *H*_1_, *H*_2_}, *c*_0_ corresponds to the global average, *age*_*i*_ is the age in years of subject *i, isFemale*_*i*_ is a binary indicator variable for the sex of subject *i* (1=female, 0=male), and *icv*_*i*_ denotes the intracranial volume of subject *i*. We assumed that the log-Jacobian of the surface transformation depends linearly on age. We also assumed that the measurement noise *ϵ*_*i*_ is independently and identically distributed zero-mean Gaussian with unknown, common variance. The unknown effects {*c*_0_, *c*_1_, *c*_2_, *c*_3_, *c*_4_, *c*_5_} were estimated via leastsquares. To determine whether the study sample is stable and that our individual results are not driven by outliers or extreme values, we performed permutation testing according to an established procedure [43]. The vestibular variable was permuted across all clusters on a surface under the null hypotheses, *H*_0*A*_ and *H*_0*B*_, for 10,000 simulations. The maximum test statistic, calculated as the maximum of the ratio of maximum squared errors of the null to the alternative model, was calculated for both the real and simulated models. The overall permutation p-value, *p*_*perm*_, is calculated as the proportion of simulated max test statistics greater than true (non-simulated) max test statistics. We rejected the null hypothesis if *p*_*perm*_ < 0.05. Thus, the p-values from testing across clusters are corrected for Family-Wise Error Rate (FWER) at the 0.05-level. Furthermore, a cluster *k* is significant if the true test statistic is greater than the 95th percentile of simulated test statistics. For clusters which rejected the null hypotheses, *H*_0*A*_ (*H*_0*B*_), 95% confidence intervals were calculated by bootstrapping model residuals under the alternative hypothesis, *H*_1_ (*H*_2_), to mitigate the effects of outliers. Bootstrapped studentized confidence intervals were computed by bootstrapping model residuals with 10,000 simulations using the *bootci* function in Matlab. All analyses were implemented in Matlab.

## 3. Results

### 3.1 Characteristics of the study sample

Table 1 shows the characteristics for the study sample from the BLSA. Two-sided t-tests show that bivariate partial correlations of vestibular function and vision/proprioception function are insignificant (*p* < 0.05) while controlling for age (Table 2). Additionally, the bivariate correlation between hearing and vestibular functions was significant while controlling for age (*p* = 0.042), but fell below significance when additionally controlling for gender (*ρ* = −0.19 (*p* = 0.066)).

**Table 1:** Characteristics of the study sample, presented on their original scale (N = 117). Key: PTA: four-frequency (0.5, 1, 2, 4 kHz) pure tone average from the better ear; n: the number of participants with a visit where both the characteristic and MRI data were available; %: 100(n/N) percent; SD: standard deviation.

| Characteristic | Mean (SD) | N (%) |
| --- | --- | --- |
| Age (years) | 77 (8.7) |  |
| Sex |  |  |
| Male |  | 79 (67.5) |
| Female |  | 38 (32.5) |
| Education (Years) | 17.1 (2.5) |  |
| Best Corrected cVEMP Amplitude | 1.2 (0.75) |  |
| Best oVEMP Amplitude (μV) | 13.6 (10.1) | |
| Mean VOR Gain | 0.997 (0.16) |  |
| Best Four Frequency PTA (dB) | 32 (14.9) |  |
| Visual Acuity (logMAR) | 0.11 (0.13) |  |
| Proprioception Threshold (degrees) | 1.71 (1.73) |  |

**Table 2:**
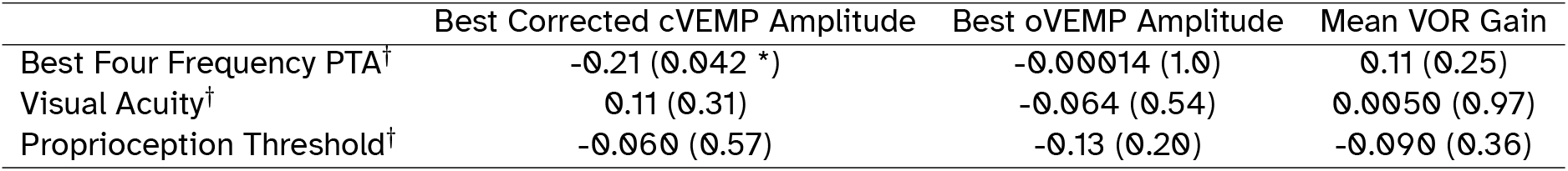
Bivariate Pearson partial correlation coefficients, with p-values in parentheses, between vestibular and multi-sensory function (N = 117). Key: PTA: four-frequency (0.5, 1, 2, 4 kHz) pure tone average from the better ear; †: Variables marked with a dagger (†) have been negated such that increasing values indicate better function; *: p < 0.05.

|  | Best Corrected cVEMP Amplitude | Best oVEMP Amplitude | Mean VOR Gain |
| --- | --- | --- | --- |
| Best Four Frequency PTA† | -0.21 (0.042 *) | -0.00014 (1.0) | 0.11 (0.25) |
| Visual Acuity† | 0.11 (0.31) | -0.064 (0.54) | 0.0050 (0.97) |
| Proprioception Threshold† | -0.060 (0.57) | -0.13 (0.20) | -0.090 (0.36) |

### 3.2 Vestibular effects on prefrontal and sensorimotor cortex morphology

Figures 3, 4, and 5 illustrate the spatial distribution of the significant vestibular-only effects from the alternative hypothesis *H*_1_ and of the significant vestibular effects from the alternative hypothesis *H*_2_ which additionally covaried for hearing, vision, or proprioception function. Adding hearing function to the model reduced the saccular and canal, but not utricular, function model sample sizes from 95 and 107 subjects to 94 and 106 subjects, respectively. Adding vision function reduced the saccular, utricular, and canal function model sample sizes from 95, 100, and 107 subjects to 90, 95, and 100 subjects, respectively. The addition of proprioception function reduced the saccular and canal, but not utricular, function model sample sizes from 95 and 107 subjects to 94 and 106 subjects, respectively.

**Figure 3.**
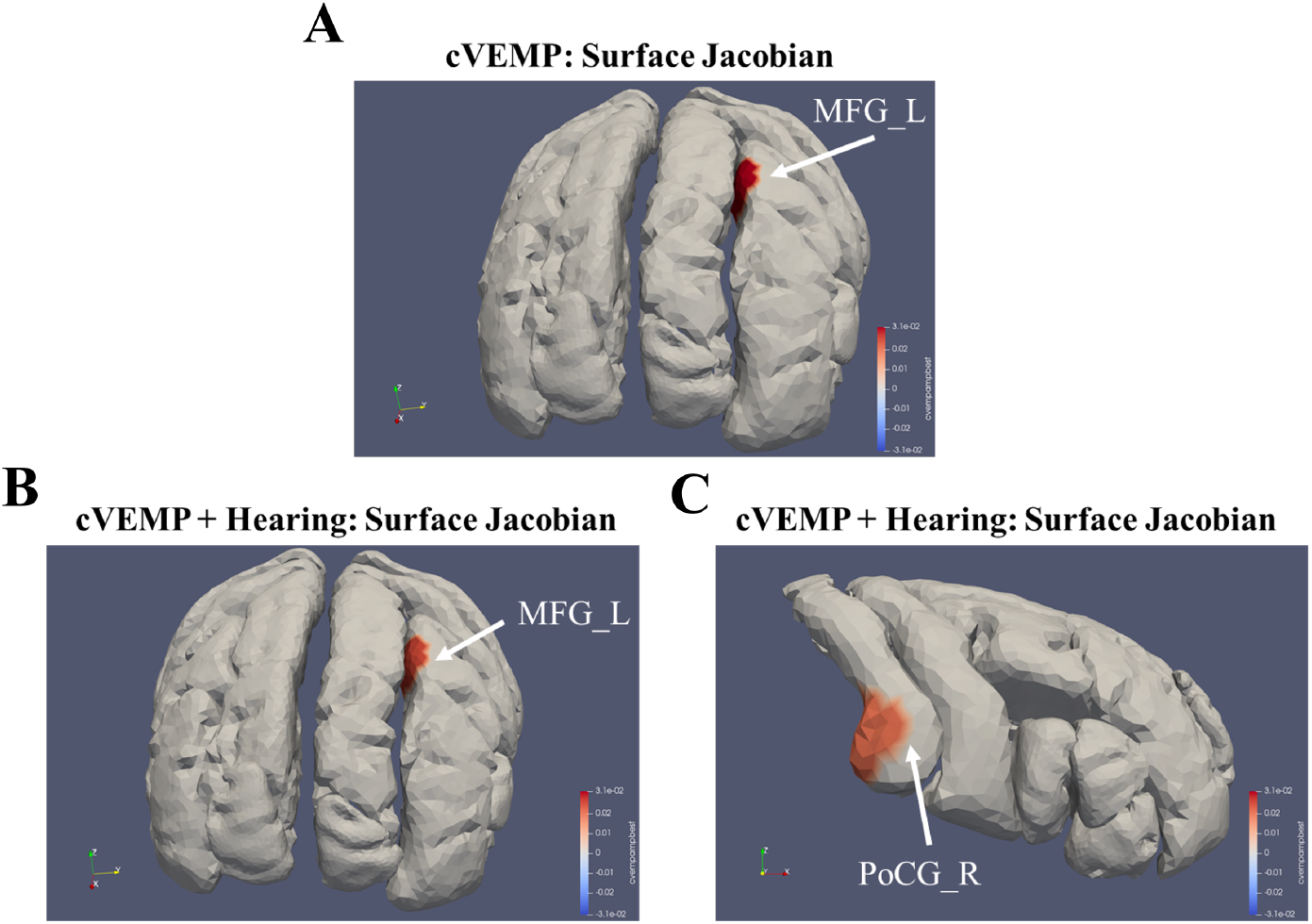
Spatial distribution of the significant saccular effects on the shapes of the frontal and sensorimotor cortices visualized on the population template. (A) shows the saccular-only results, and (B,C) show the saccular-hearing results. Regions that are colored red (blue) indicate a significant surface expansion (compression) in the direction tangent to the surface (surface Jacobian) with higher saccular function. *MFG*, posterior pars of middle frontal gyrus, *PoCG*, postcentral gyrus of the sensorimotor cortex.

**Figure 4.**
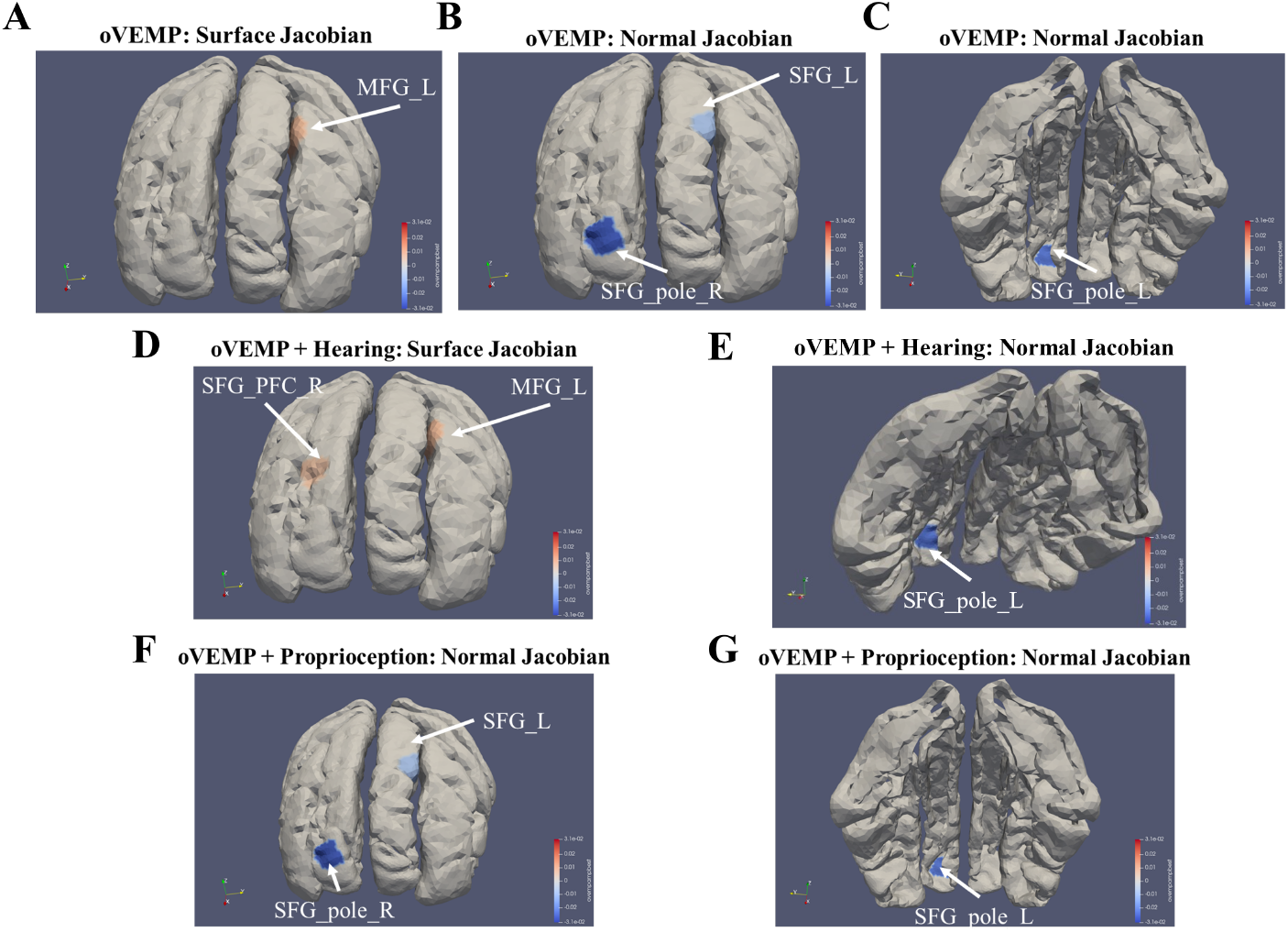
Spatial distribution of the significant utricular effects on frontal cortex shape visualized on the population template. (A-C) show the utricular-only results, (D,E) show the utricular-hearing results, and (F,G) show the utricular-proprioception results. Red (blue) indicates a region of significant surface expansion (compression) in the direction tangent/normal to the surface (surface/normal Jacobian) with higher utricular function. *MFG*, posterior pars of middle frontal gyrus, *SFG_PFC*, the middle-superior part of the prefrontal cortex, *SFG_pole*, frontal pole, and *SFG*, posterior pars of the superior frontal gyrus.

**Figure 5.**
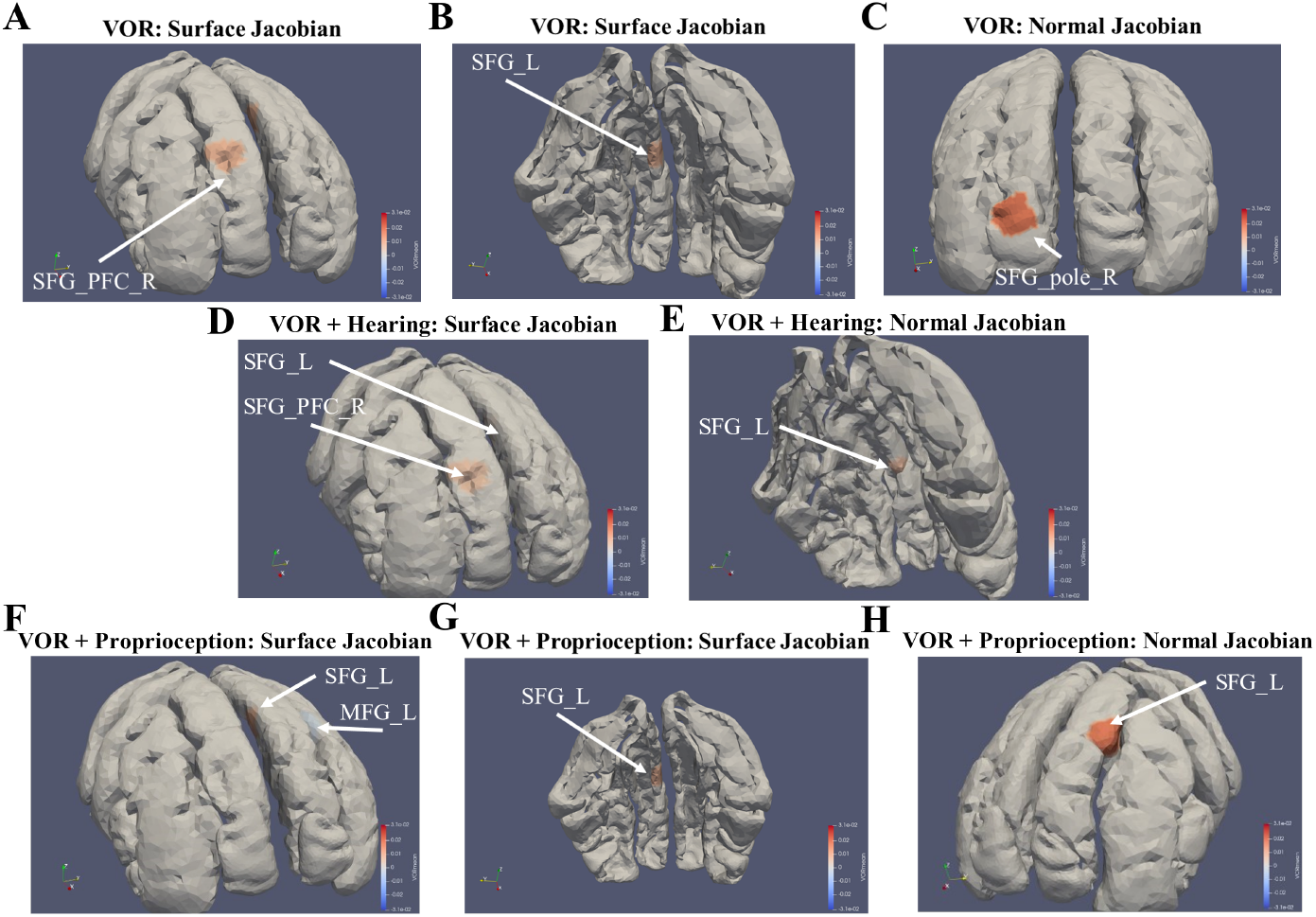
Spatial distribution of the significant horizontal canal effects on frontal cortex shape visualized on the population template. (A-C) show the utricular-only results, (D,E) show the utricular-hearing results, and (F-H) show the utricular-proprioception results. Red (blue) indicates a region of significant surface expansion (compression) in the direction tangent/normal to the surface (surface/normal Jacobian) with higher canal function. *MFG*, posterior pars of middle frontal gyrus, *SFG_PFC*, the middle-superior part of the prefrontal cortex, *SFG_pole*, frontal pole, and *SFG*, posterior pars of the superior frontal gyrus.

#### 3.2.1 Prefrontal cortex

In the vestibular-only analyses, several relationships between vestibular end-organ function and surface shape alterations in the prefrontal cortex were significant according to permutation testing. A 1 standard deviation (SD) increase in saccular function was associated with a 0.031% expansion tangent to the cortical surface in the medial left posterior MFG (*p* ≈ 0.04, CI: (−0.028, 0.091)). A 1SD increase in utricular function was associated with a 0.008% expansion tangent to the cortical surface in the medial left posterior MFG (*p* ≈ 0.018, CI: (−0.048, 0.064)), a 0.009% compression normal to the cortical surface in the rostral lateral region of the left SFG (*p* ≈ 0.047, CI: (−0.041, 0.023)), a 0.023% compression normal to the cortical surface in the caudal dorsal region of the left SFG_pole (*p* ≈ 0.0019, CI: (−0.054, 0.007)), and a 0.027% compression normal to the cortical surface in the dorsal region of the right SFG_pole (*p* ≈ 0.031, CI: (−0.060, 0.008)). A 1SD increase in canal function was associated with a 0.008% expansion tangent to the cortical surface in the medial rostral region of the left SFG (*p* ≈ 0.034, CI: (−0.033, 0.050)), a 0.008% expansion tangent to the cortical surface in the dorsal lateral region of the right SFG_PFC (*p* ≈ 0.042, CI: (−0.037, 0.054)), and a 0.018% expansion normal to the cortical surface in the dorsal region of the right SFG_pole (*p* ≈ 0.035, CI: (−0.014, 0.052)). Figures 3, 4, and 5 show the spatial distribution of these associations.

In the multi-sensory analyses of otolith function, many relationships persisted and others were found (See Figures 3, 4, and Supplementary Table 1). Figure 3B shows a 1SD increase in saccular function was associated with a 0.025% expansion tangent to the cortical surface in the medial left posterior MFG after accounting for hearing function (*p* ≈ 0.0283, CI: (−0.007, 0.11)). A 1SD increase in saccular function was associated with a 0.025% expansion tangent to the cortical surface in the medial left posterior MFG after accounting for hearing function (*p* ≈ 0.0283, CI: (−0.007, 0.11)) (See Figure 3B). In models additionally covarying for hearing function, a 1SD increase in utricular function was associated with a 0.008% expansion tangent to the cortical surface in the medial left MFG (*p* ≈ 0.0164, CI: (−0.038, 0.069)), a 0.006% expansion tangent to the the dorsal lateral surface of the right SFG_PFC (*p* ≈ 0.0157, CI: (−0.055, 0.025)) (Figure 4D), and a 0.023% compression normal to the caudal surface of the left SFG_pole (*p* ≈ 0.0025, CI: (−0.021, 0.045)) (Figure 4D). In models additionally covarying for proprioceptive function, a 1SD increase in utricular function was associated with a 0.014% compression normal to the cortical surface in the rostral lateral left SFG (*p* ≈ 0.0312, CI: (0.027, 0.105)), a 0.025% compression normal to the cortical surface in the rostral dorsal left SFG_pole (*p* ≈ 0.0025, CI: (−0.026, 0.041)), and a 0.031% compression normal to the caudal surface of left SFG_pole (*p* ≈ 0.037, CI: (−0.096, -0.012)) (See Figures 4F and 4G).

In the multi-sensory analyses of canal function, many relationships persisted and numerous others were found (See Figure 5 and Supplementary Table 1). Figures 5D and 5E show a 1SD increase in canal function was associated with a 0.002% expansion tangent to the cortical surface in the medial rostral region of the left SFG (*p* ≈ 0.035, CI: (−0.043, 0.044)), a 0.007% expansion normal to the cortical surface in the medial rostral region of the left SFG (*p* ≈ 0.037, CI: (−0.02, 0.033)), and a 0.006% expansion tangent to the cortical surface in the dorsal lateral region of the right SFG_PFC (*p* ≈ 0.014, CI: (−0.042, 0.052)), after accounting for hearing function. In the canal-proprioception function models, a 1SD increase in canal function was associated with a 0.003% compression tangent to the cortical surface in the medial left posterior MFG, a 0.008% expansion tangent to the cortical surface in the medial rostral region of the left SFG (*p* ≈ 0.033, CI: (−0.025, 0.051)), and a 0.018% expansion normal to the cortical surface in the medial rostral region of the left SFG (*p* ≈ 0.027, CI: (−0.011, 0.047)) (See Figures 5F, 5G, and 5H).

No relationships between vestibular function and the shape of the inferior frontal gyrus (pars opercularis, pars triangularis, pars orbitalis), or the MFG_DPFC survived permutation testing at the 0.05 level. Notably, these findings persist and more are uncovered after accounting for hearing function, vision function, and proprioceptive function in individual bivariate analyses (See Figures 3, 4, and 5 and Supplementary Table 1). Furthermore, all relationships between vestibular function and the shapes of the frontal and sensorimotor cortices were attenuated when correcting for vision function, but many relationships still showed strong trends in the left MFG (canal function: *p* ≈ 0.066), left SFG (canal function: *p* ≈ 0.056), and right SFG_PFC (canal function: *p* ≈ 0.087). Moreover, we note several strong trends in the multisensory analyses that did not survive permutation testing at the 0.05 level (See Supplementary Table 1).

#### 3.2.2 Sensorimotor cortex

No relationships between vestibular function and the shape of the precentral gyrus or the postcentral gyrus survived permutation testing at the 0.05 level (i.e. all *p*_*perm*_ ≥ 0.05) in the nonmulti-sensory analysis. Importantly, a relationship was found after accounting for hearing function, but not vision or proprioceptive function in separate bivariate analyses (See Figure 3C and Supplementary Table 1). Permutation testing revealed a significant relationship between saccular function and tangent surface shape in the right poCG when additionally covarying for hearing function. Figure 3C shows that a 1SD increase in saccular function correlated with approximately 0.020% expansion tangent to the cortical surface in the posterior ventrolateral surface of the right PoCG (*p* ≈ 0.0242, CI: (−0.075, 0.010)). Despite not surviving permutation testing at the 0.05 level, there were several strong trends in the multisensory analyses, in particular in the left PoCG (canal-vision function model: *p* ≈ 0.079), left PrCG (saccular-vision function model: *p* ≈ 0.091), and right PrCG (canal-proprioception function model: *p* ≈ 0.1) (See Supplementary Table 1).

## 4 Discussion

In this study of healthy, older adults, we found that reduced vestibular function is associated with shape alterations in ten ROIs of the putative prefrontal and sensorimotor “vestibular cortex”. The ROIs investigated include the middle-superior part of the prefrontal cortex (SFG_PFC), frontal pole (SFG_pole), and posterior pars of the superior frontal gyrus (SFG), the dorsal prefrontal cortex and posterior pars of middle frontal gyrus (MFG_DPFC, MFG), the pars opercularis, pars triangularis, and pars orbitalis of the inferior frontal gyrus (IFG), as well as the precentral gyrus (PrCG) and postcentral gyrus (PoCG) of the sensorimotor cortex. Specifically, we found associations between reduced saccular function and significant cortical surface compression in the MFG, reduced utricular function and MFG compression and expansion of the SFG and SFG_pole, respectively, and reduced canal function and surface compression of the SFG, SFG_PFC, and SFG_pole. After additionally adjusting for measures of hearing and proprioception, we observed shape alterations in the MFG, SFG, SFG_PFC, SFG_pole, and PoCG with poorer end-organ functions. However, additionally adjusting for vision function attenuated the observed relationships, albeit they exhibited strong trends toward significance. This finding likely stems from a power loss resulting from a redistribution of explained variance, thereby reducing the vestibular-only effect size. Additionally, the loss of power is likely influenced by a small reduction in degrees of freedom (e.g. adding vision reduced the saccular, utricular, and canal function model sample sizes from 95, 100 and 107 subjects to 90, 95 and 100 subjects, respectively). This loss of power raises the detectable vestibular-only effect size for our sample size, and thus a larger sample size would be needed to detect vestibular effects in the presence of vision effects. Furthermore, a ceiling/floor effect of vision function could lead to overestimation of the vision effect, further exacerbating the issue with the detectable vestibular effect size. Importantly, given that vestibular and vision functions were insignificantly correlated, and a larger sample size would allow the vestibular-only effects to be revealed in the presence of vision function, we suspect that the relationship between vestibular function and local frontal cortex morphology is independent of vision function. The significant structures are known to exhibit robust activations to artificial and naturalistic vestibular stimulation as well as structural alterations in aging and vestibular syndromes [66, 67, 68, 41, 34, 37, 39, 42]. Our findings align with previous links between vestibular function and the structures of the somatosensory [34, 41, 42], motor [39, 41], and prefrontal cortices [37, 39, 41, 42] and clarify previous inconsistent reports [34, 35, 36, 37, 38, 39, 40, 41, 42].

### 4.1 Prefrontal cortex

Our findings support the initial hypothesis that diminished vestibular function correlates with structural changes in the prefrontal cortex, largely independent of multisensory functions. Specifically, reductions in saccular and utricular functions are associated with surface compression in the medial left MFG, irrespective of auditory function. Additionally, decreased canal function correlates with compressions in the medial rostral region of the left SFG, independent of both hearing and proprioception functions, and in the dorsal lateral region of the right SFG_PFC, independent of hearing function alone. In the context of age-related auditory changes, reduced canal function is associated with compression in the most rostral medial region of the left SFG. Conversely, considering age-related proprioceptive changes, reduced canal function is linked to both an expansion in the rostral medial surface of the left MFG and a compression in the rostral lateral surface of the left SFG. Interestingly, the compressive effect on the dorsolateral surface of the right SFG_PFC, associated with diminished utricular function, is insignificant when accounting for age-related hearing changes. Unexpectedly, reduced utricular function is also associated with expansions in the rostral lateral region of the left SFG, the caudal dorsal region of the left SFG_pole, and the rostral dorsal region of the right SFG_pole, independent of proprioceptive functions. These expansions, which may reflect age-related alterations in vestibular sensitivity, are moderated by auditory function, as evidenced by attenuations in the expansions of the right SFG_pole and left SFG [69, 70].

The MFG and SFG, which contain the premotor cortex, supplementary motor area, and frontal eye fields, are crucial for motor control, planning, and initiating visuospatial movements. These regions are interconnected with various brain regions, including other prefrontal areas, premotor, cingulate, somatosensory, and insular regions, facilitating the coordination of working memory for actions and complex planning sequences [71]. Vestibular inputs to these areas, supported by animal and human studies, suggest significant vestibular influence on regions near the frontal eye fields and the supplementary motor area [1, 72, 73]. Evidence from subclinical and clinical studies further suggests that vestibular impairments correlate with structural changes in these cortical areas, emphasizing their role in vestibular processing [41, 37, 39]. Moreover, the frontal pole’s extensive connectivity and role in episodic memory (lateral subregion) and cognitive switching (anterior medial subregion) suggest its critical involvement in managing deficits in cognitive-motor dual-tasking observed in vestibular patients [74, 75, 76, 77]. Despite no detected relationship in the literature at the time of writing between dual-task gait performance and the SFG_pole [78], we speculate that the frontal pole utilizes utricle-and canal-transduced linear and angular acceleration data to coordinate complex body and visuo-motor actions (e.g. locomotion, reaching, and grasping) while balancing various cognitive, interoceptive, and emotional demands.

Unexpectedly, no significant correlations were found between vestibular function and the shapes of the pars opercularis, pars orbitalis, and pars triangularis of the inferior frontal gyrus, or the MFG_DPFC. This absence of expected correlations, despite extensive literature highlighting the involvement of these areas in vestibular processing [66], suggests two possible explanations. The first is that we may be more likely to detect volume-based, or cortical thickness-based changes in these regions. The second is that there are potential compensatory mechanisms within the brain that adjust to age-related changes in vestibular sensitivity [69, 70]. Furthermore, the peripheral vestibular information processed through thalamic-limbic-striatal-frontal circuits likely integrates with multi-sensorimotor data before reaching the prefrontal cortex. This integration could explain the lack of observed structural changes as compensatory adaptations or differential sensitivities to combined sensory inputs, influenced by aging and neuroplasticity. Together, an age-related and multi-sensory involvement could explain how these regions respond to otolith or canal information and show no relationship with structural alterations (and language function [79, 80]) in older adults. Overall, our results underscore a significant link between age-related declines in vestibular function and morphological variations in the frontal cortex, suggesting a broader impact of sensory integration on cognitive and motor functions in older adults [29, 81, 82, 83].

### 4.2 Sensorimotor cortex

It is unclear why, in the vestibular-only analysis, there were no associations of saccular, utricular, or horizontal semi-circular canal function with the PrCG or PoCG in either hemisphere, given these regions are implicated in the vestibular cognitive network [6, 1, 5, 3]. It may be the case that we are more likely to detect changes in volume or thickness in these regions. However, in the multi-sensory analysis, reduced saccular function correlated with compression in the posterior ventrolateral region of the right PoCG when accounting for age-related hearing loss. Previous studies of vestibular stimulation reported robust neural responses in the primary and secondary somatosensory cortex in rats [84] and in humans [1]. Because the posterior ventrolateral region of the right PoCG may contain the representation of the mouth and larynx, it is connected with the multi-sensorimotor speech and language network (e.g. the supramarginal gyrus), and speech activates both the auditory and vestibular systems [85, 86, 87], this finding may imply the importance of saccular and hearing function in the context of speech planning and execution. We speculate that this finding may also be important for self-other voice discrimination, which relies on auditory, somatosensory (e.g. bone-conducted vibration signals, mouth proprioception), and vestibular processing [86, 87] by the PoCG. Additionally, the connectivity of the posterior ventrolateral region of the PoCG, approximately corresponding to BA 1 and BA 2, with key vestibular network regions (e.g. the insula) implies an important role more broadly in somatosensation, bodily self-consciousness and control, and motor planning which involves self-motion perception, and social cognition [32, 33, 71, 88, 89].

Whether the PoCG may use information about linear acceleration of the head in the horizontal plane transduced by the utricle or about angular acceleration of the head in the horizontal plane (yaw) transduced by the horizontal semi-circular canal must be elucidated. Older adults with reduced vestibular function as measured by the standing on foam with eyes closed balance (FOEC) test (i.e. more sway) were observed to have poorer sensorimotor cortex structure [41]. This is important as one study of older adults found that age-related horizontal canal dysfunction is associated with decreased performance on the FOEC test [90]. Additionally, older adults were observed to have significantly shallower sulcal depth (i.e. worse brain structure) in the the sensorimotor, supramarginal, insular, and superior frontal and parietal cortices with poorer dual-gait performance [78].

### 4.3 Strengths of this study

We report several strengths of this study. One such strength is that the relationships examined were hypotheses-driven based on converging evidence from structural and functional neuroimaging in humans. A second strength is that we use a state-of-the-art brain mapping pipeline. This pipeline utilizes a study-appropriate multi-atlas and LDDMM, a well-established framework of non-linear image registration techniques [91]. Moreover, we employed LDDMM-based surface diffeomorphometry to overcome the limitations encountered by other vestibular neuroimaging studies that used low-strength MRI, voxel-based morphometry, or volume-based morphometry, such as the tendency to miss effects that are subtle, non-focal, or non-uniformly spatially distributed across the region of interest [92]. Surface diffeomorphometry provides a sensitive measure of cortical shape variation and has been used to track sub-voxel structural alterations in aging and disease [43, 55, 56, 57, 58, 59, 60, 61]. A third strength is that our quality control pipeline involved manual inspections of the data at each step of processing. Also, our statistical testing pipeline accounts for multiple comparisons as well as for outliers using permutation testing and bootstrapping, respectively. In contrast to vestibular neuroimaging studies that stimulate the end-organs in a combination of ways (e.g. galvanic vestibular stimulation; caloric stimulation), we use individual measurements of the utricle, saccule, and horizontal semi-circular canal to capture end-organ specific relationships with brain morphology. This is important for aging studies because the hair cells in the cristae of the semi-circular canals decline with age earlier than those of the otolithic maculae. Thus, their individual contributions to the aging of the central vestibular pathways may be different. We also used specific clinical assessments of hearing, vision, and proprioception function to determine multi-sensory involvement, rather than use a composite clinical test based on gait or balance/posturography.

### 4.4 Limitations of this study

We note several limitations to this study. While cortical surface shape analysis provides sensitive measures morphology, it only describes how the surface is altered. Thus, structural changes within the structure of interest are missed. While volume measures the size of the broader structure and complements the local shape measures when the changes are uniform across (not within) the structure, cortical thickness may provide a complementary sensitive measure of cortical morphology, and when paired with equivolumetric theory, can describe what happens within each structure of interest in terms of layer thicknesses [93]. The normal Jacobian measure used in this study is qualitatively different than a cortical thickness measure, which is often defined as the length of a streamline connecting two opposing points on the cortical ribbon. Recent studies have highlighted the importance of cortical microarchitecture and layer-specific relationships in cognitive networks [94]. The small local changes in shape could be compensated for by shape changes in the other direction in the rest of the broader structure (even if the latter changes are each non-significant). This has been shown before in studies of children with ADHD in whom basal ganglia volume changes in some cases conflicted with the direction of local shape changes [95, 96].

Notably, the reproducibility of findings is a challenge for several reasons. Anatomical definitions can vary between atlases and experts, adding to the great variability in appearance of cortical parcels at high granularity. The quality and smoothness of surface triangulations, as well as the choice of surface mapping algorithm parameters, also may impact the results. The surface clustering approach may impact the results. The choice of number of surface clusters determines the spatial extent of the cluster, which implicitly corresponds to an assumption about the size of the region related to the effect. In this study, we chose the number of clusters to balance the number of patches, and therefore the number of comparisons, and the spatial extent of the effects. To consider a continuum of surface cluster sizes, a threshold-free cluster enhancement procedure for surfaces could be developed. Additionally, the boundaries of the clusters may overlap regions where the true effect lies and noise in such a way to mask the true effect. Moreover, our population templates are created based on this particular sample; thus, they would be different when creating a new population template based on a different sample. Although the test statistic we used was based on maximum squared errors, which is generally less robust than the sum of squared errors, it is a conservative approach typically employed in our group. This approach is conservative because it compares our results to the least favorable outcome, rather than average outcomes, under permuted vestibular function. Due to our nonparametric testing procedure, the validity of our p-values is independent of the data distribution and of our choice of test statistic, whereas the power of the tests is dependent.

Although our large multi-atlas set spans our investigated age range to capture the anatomical variability of adult brains, this multi-atlas set lacks modern cytoarchitectonic definitions that have relevance to brain function. Another limitation is that we did not examine the potential role of the cerebellum, the brainstem, the hypothalamus, or the thalamus in modulating the effects of age-related vestibular loss in the cortex. However, robust measures of cerebellar and brainstem structures are being developed. While we did reuse data, we did not account for dataset decay [97] because we explored a distinct research question compared to previous studies from our group that use this cohort and measurements [43, 44]. This may limit the strength of our findings, and future confirmatory studies may be needed to reinforce our findings. Additionally, our findings may not generalize to the broader and younger population due to the age range used in this study and the propensity of BLSA participants to have higher levels of education and socioeconomic status than typical adults. These are important caveats to the interpretation of our findings, as higher education and socioeconomic status may be associated with frontal and sensorimotor structure. Finally, these results may not generalize to younger adults who have weakened vestibular function.

### 4.5 Future work

To understand the neuroanatomical underpinnings of aging on vestibular-mediated behaviors, several studies will be needed. Longitudinal studies incorporating gray matter volume, shape, and cortical thickness and white matter microstructural integrity of the limbic system, temporoparietal junction, and frontal cortex will help to understand the relationships over time. Because the brain vestibular network is plastic and compensates for vestibular loss to maintain behavioral function, we aim to use changepoint analysis to identify subtle non-linearities in the trends of brain structure alterations that may be missed by gross aging trends [98]. Then a precedence graph can be created that highlights the sequence of regional structural changes in relation to each other. To investigate causal hypotheses between vestibular loss and structural changes in the multi-sensorimotor vestibular network, our group plans to use longitudinal structural equation modeling that accounts for possible confounding by multi-sensorimotor function. Notably, structural equation modeling can test hypotheses regarding the relationship between vestibular-mediated behaviors and intermediating brain regions (e.g. brainstem, hypothalamus, cerebellum, thalamus). By harmonizing our atlas definitions with modern brain atlases based on cytoarchitecture [99, 100] or multi-modal parcellations [100, 101, 102], new insights into structure-function relationships can be gleaned. Altogether, this future work can reveal the sequence and causal direction of changes in the multi-sensorimotor vestibular network.

## 5. Conclusion

Our findings highlight subtle associations between age-associated vestibular loss and the structure of the frontal cortex—a key region in the vestibular cognitive network that receives multi-sensorimotor vestibular information—in-line with previous neuroimaging studies of vestibular function. Furthermore, these findings may provide the neuroanatomical links between vestibular loss and higher-order cognitive deficits observed in the aging population and in people with dementia or Parkinson’s disease. Future work will need to determine the temporal and spatial flow of structural alterations in brain regions that receive vestibular information and that are involved in vestibular-mediated behaviors, such as self-motion perception, motor planning, and executive function. Bolstering the understanding of the involvement of peripheral and central vestibular loss in self-motion perception, motor planning, and executive function will be vital for the development of sensible interventions.

## Data Availability

The BLSA data are available upon request on the BLSA website (blsa.nih.gov). Requests undergo a review by the BLSA Data Sharing Proposal Review Committee and approval from the NIH Institutional Review Board.

https://www.blsa.nih.gov

## Acknowledgments

This work was supported by the National Institute on Aging (Grant R01 AG057667), the National Institute on Deafness and Other Communication Disorders (Grant R03 DC015583), and the National Institute of Biomedical Imaging and Bioengineering (Grant P41-EB031771).

## Supporting Information

See the Supplementary Material.

## 6 Supplementary Material

**Supplementary Table 1:**
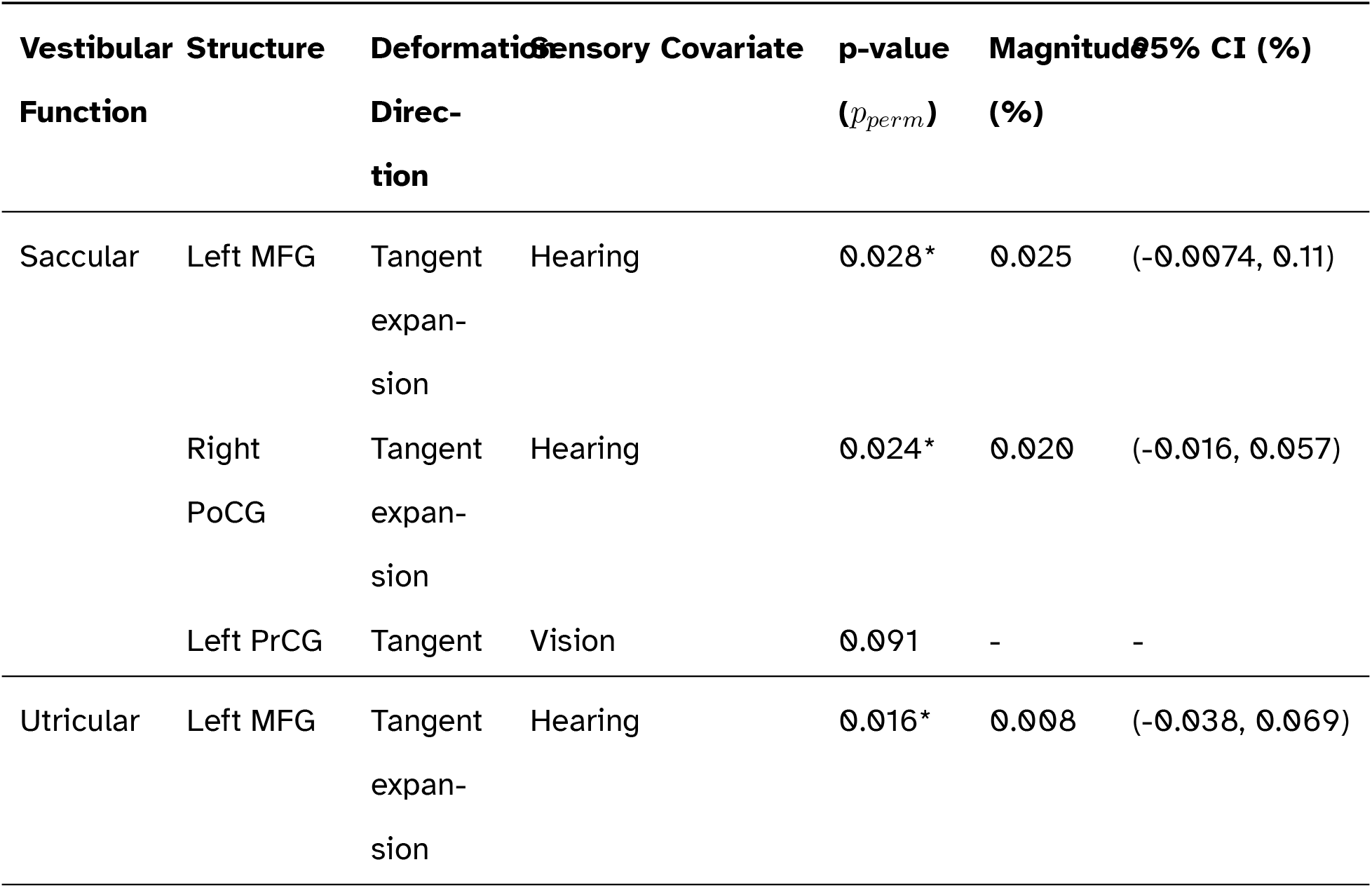
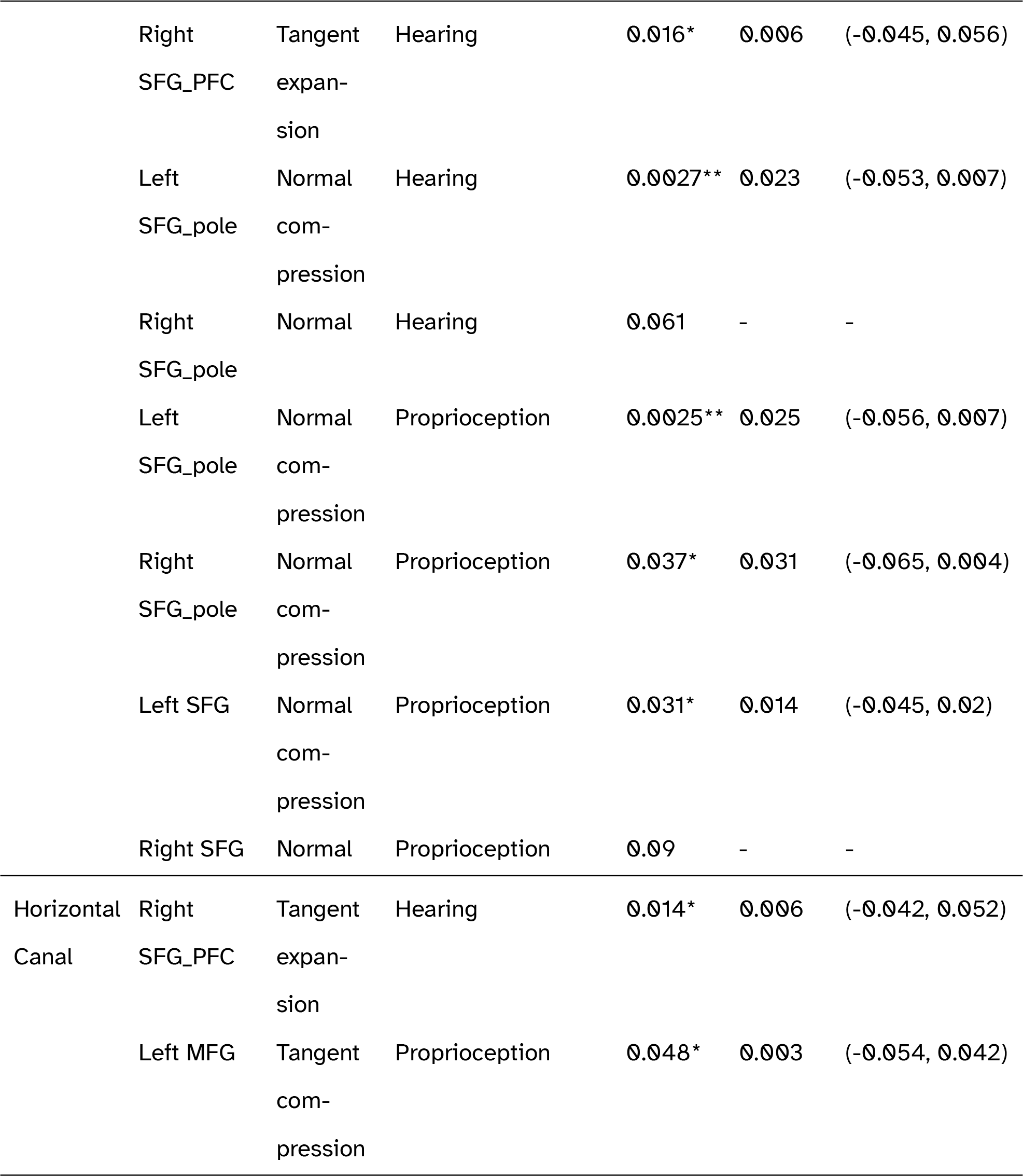
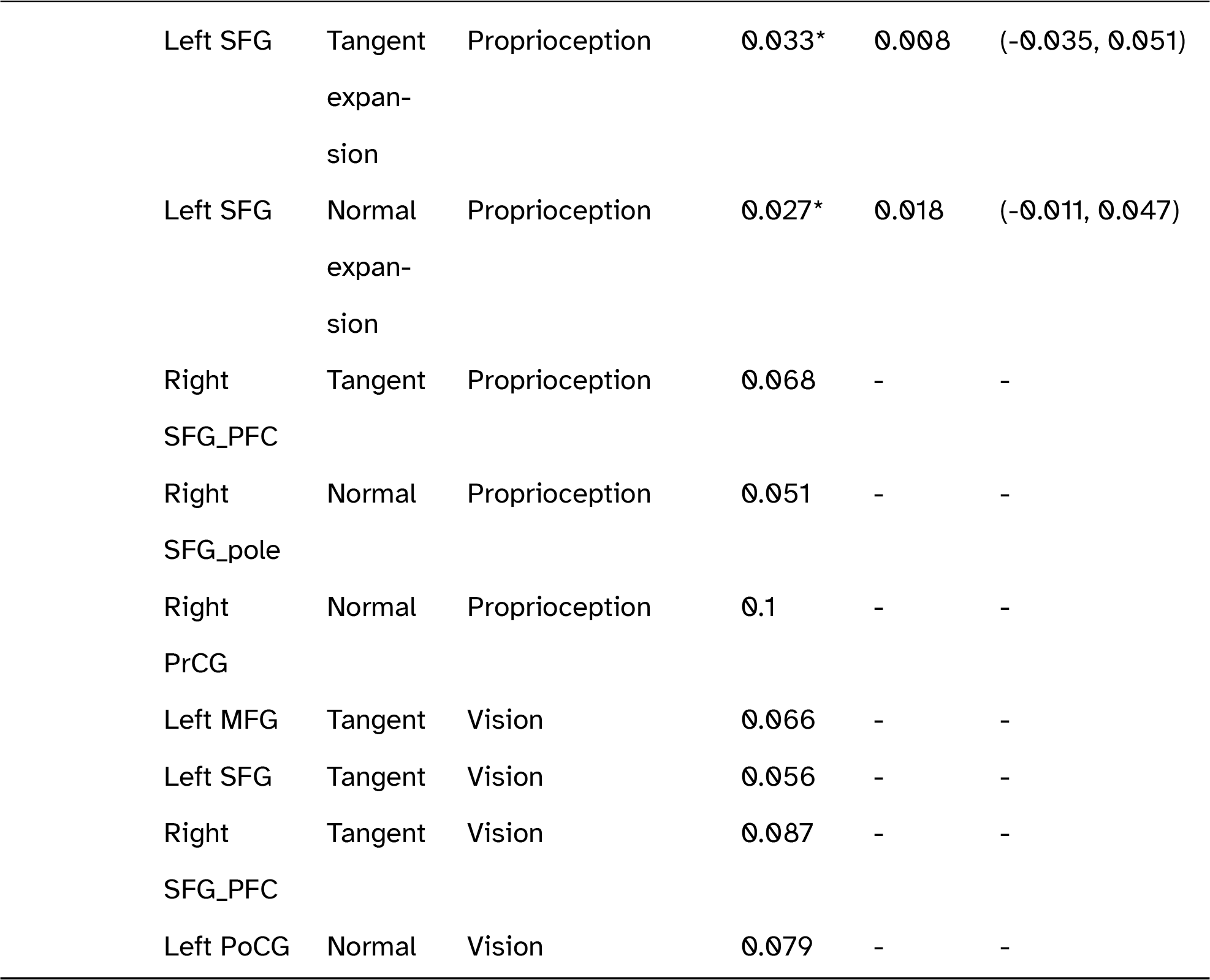
Significant and strongly trending results of the multisensory regression models, in which hearing, vision, and proprioception function were included as covariates. A tangent expansion (compression) corresponds to a positive (negative) log-surface Jacobian, and similarly for a normal expansion (compression) and a positive (negative) log-normal Jacobian. Key: MFG: posterior middle frontal gyrus; SFG: posterior superior frontal gyrus; SFG_PFC: prefrontal cortex part of the superior frontal gyrus; SFG_pole: frontal pole of the superior frontal gyrus; PrCG: precentral gyrus; PoCG: postcentral gyrus; CI: confidence interval; **\*\* p<0.01, * p<0.05**.

| Vestibular Function | Structure | Deformation Direction | Sensory Covariate | p-value ($p_{perm}$) | Magnitude (%) | 95% CI (%) |
| --- | --- | --- | --- | --- | --- | --- |
| Saccular | Left MFG | Tangent expansion | Hearing | 0.028* | 0.025 | (-0.0074, 0.11) |
|  | Right PoCG | Tangent expansion | Hearing | 0.024* | 0.020 | (-0.016, 0.057) |
|  | Left PrCG | Tangent | Vision | 0.091 | - | - |
| Utricular | Left MFG | Tangent expansion | Hearing | 0.016* | 0.008 | (-0.038, 0.069) |
|  | Right SFG_PFC | Tangent expansion | Hearing | 0.016* | 0.006 | (-0.045, 0.056) |
|  | Left SFG_pole | Normal compression | Hearing | 0.0027** | 0.023 | (-0.053, 0.007) |
|  | Right SFG_pole | Normal | Hearing | 0.061 | - | - |
|  | Left SFG_pole | Normal compression | Proprioception | 0.0025** | 0.025 | (-0.056, 0.007) |
|  | Right SFG_pole | Normal compression | Proprioception | 0.037* | 0.031 | (-0.065, 0.004) |
|  | Left SFG | Normal compression | Proprioception | 0.031* | 0.014 | (-0.045, 0.02) |
|  | Right SFG | Normal | Proprioception | 0.09 | - | - |
| Horizontal Canal | Right SFG_PFC | Tangent expansion | Hearing | 0.014* | 0.006 | (-0.042, 0.052) |
|  | Left MFG | Tangent compression | Proprioception | 0.048* | 0.003 | (-0.054, 0.042) |
|  | Left SFG | Tangent expansion | Proprioception | 0.033* | 0.008 | (-0.035, 0.051) |
|  | Left SFG | Normal expansion | Proprioception | 0.027* | 0.018 | (-0.011, 0.047) |
|  | Right SFG_PFC | Tangent | Proprioception | 0.068 | - | - |
|  | Right SFG_pole | Normal | Proprioception | 0.051 | - | - |
|  | Right PrCG | Normal | Proprioception | 0.1 | - | - |
|  | Left MFG | Tangent | Vision | 0.066 | - | - |
|  | Left SFG | Tangent | Vision | 0.056 | - | - |
|  | Right SFG_PFC | Tangent | Vision | 0.087 | - | - |
|  | Left PoCG | Normal | Vision | 0.079 | - | - |

## References

[1] Christophe Lopez and Olaf Blanke. “The thalamocortical vestibular system in animals and humans”. In: Brain Research Reviews 67.1 (June 2011), pp. 119–146. DOI: 10.1016/j.brainresrev.2010.12.002. URL: https://www.clinicalkey.es/playcontent/1-s2.0-S0165017311000026.

[2] Julian Conrad, Bernhard Baier, and Marianne Dieterich. “The role of the thalamus in the human subcortical vestibular system1”. In: Journal of Vestibular Research 24.5-6 (2014), p. 375. DOI: 10.3233/VES-140534.

[3] C. De Waele et al. “Vestibular projections in the human cortex”. In: Experimental brain research 141.4 (Dec. 2001), pp. 541–551. DOI: 10.1007/s00221-001-0894-7. URL: https://www.ncbi.nlm.nih.gov/pubmed/11810147.

[4] Ria Maxine Ruehl et al. “The human egomotion network”. In: NeuroImage 264 (2022). DOI: 10.1016/j.neuroimage.2022.119715.

[5] Martin Hitier, Stephane Besnard, and Paul F. Smith. “Vestibular pathways involved in cognition”. In: Frontiers in Integrative Neuroscience 8 (July 2014), p. 59. DOI: 10.3389/fnint.2014.00059. URL: https://www.ncbi.nlm.nih.gov/pubmed/25100954.

[6] Elisa Raffaella Ferrè and Patrick Haggard. “Vestibular cognition: State-of-the-art and future directions”. In: Cognitive neuropsychology 37.7-8 (Nov. 2020), pp. 413–420. DOI: 10.1080/02643294.2020.1736018. URL: https://www.tandfonline.com/doi/abs/10.1080/02643294.2020.1736018.

[7] Aisha Harun et al. “Vestibular Impairment in Dementia”. In: Otology & Neurotology 37.8 (2016), pp. 1137–1142. DOI: 10.1097/MAO.0000000000001157.

[8] Eric X. Wei et al. “Vestibular Loss Predicts Poorer Spatial Cognition in Patients with Alzheimer’s Disease”. In: Journal of Alzheimer’s Disease 61.3 (Jan. 2018), p. 995. DOI: 10.3233/JAD-170751.

[9] Eric X. Wei et al. “Saccular Impairment in Alzheimer’s Disease Is Associated with Driving Difficulty”. In: Dementia and Geriatric Cognitive Disorders 44.5-6 (Jan. 2019), p. 294. DOI: 10.1159/000485123.

[10] Kevin Biju et al. “Vestibular Function Predicts Balance and Fall Risk in Patients with Alzheimer’s Disease”. In: Journal of Alzheimer’s disease 86.3 (Jan. 2022), pp. 1159–1168. DOI: 10.3233/JAD-215366. URL: https://www.ncbi.nlm.nih.gov/pubmed/35180117.

[11] Graham D. Cochrane et al. “Cognitive and Central Vestibular Functions Correlate in People With Multiple Sclerosis”. In: Neurorehabilitation and Neural Repair 35.11 (2021), p. 1030. DOI: 10.1177/15459683211046268.

[12] Udo Rüb et al. “Huntington’s Disease (HD): Degeneration of Select Nuclei, Widespread Occurrence of Neuronal Nuclear and Axonal Inclusions in the Brainstem”. In: Brain Pathology 24.3 (Mar. 2014), p. 247. DOI: 10.1111/bpa.12115.

[13] Paul F Smith. “Vestibular functions and Parkinson’s disease”. In: Frontiers in neurology 9 (2018), p. 427861.

[14] Wenqi Cui, Zhenghao Duan, and Juan Feng. “Assessment of Vestibular-Evoked Myogenic Potentials in Parkinson’s Disease: A Systematic Review and Meta-Analysis”. In: Brain sciences 12.7 (July 2022), p. 956. DOI: 10.3390/brainsci12070956. URL: https://search.proquest.com/docview/2693939246.

[15] Sandra Carpinelli et al. “Distinct Vestibular Evoked Myogenic Potentials in Patients With Parkinson Disease and Progressive Supranuclear Palsy”. In: Frontiers in Neurology 11 (Feb. 2021). DOI: 10.3389/fneur.2020.598763.

[16] Nicolaas I. Bohnen et al. “Decreased vestibular efficacy contributes to abnormal balance in Parkinson’s disease”. In: Journal of the Neurological Sciences 440 (Sept. 2023). ISSN: 0022-510X. DOI: 10.1016/j.jns.2022.120357.

[17] Güler Berkiten et al. “Assessment of the Clinical Use of Vestibular Evoked Myogenic Potentials and the Video Head Impulse Test in the Diagnosis of Early-Stage Parkinson’s Disease”. In: Annals of Otology, Rhinology & Laryngology 132.1 (2023), pp. 41–49.

[18] Jeong-Ho Park and Suk Yun Kang. “Dizziness in Parkinson’s disease patients is associated with vestibular function”. In: Scientific Reports 11.1 (Sept 2021). DOI: 10.1038/s41598-021-98540-5.

[19] Jeong Ho Park, Min Seung Kim, and Suk Yun Kang. “Initial Vestibular Function May Be Associated with Future Postural Instability in Parkinson’s Disease”. In: Journal of Clinical Medicine 11.19 (Sept 2022). DOI: 10.3390/jcm11195608.

[20] Nathalie Chastan et al. “Prediagnostic markers of idiopathic Parkinson’s disease: Gait, visuospatial ability and executive function”. In: Gait & Posture 68 (Feb. 2019), p. 500. ISSN: 0966-6362. DOI: 10.1016/j.gaitpost.2018.12.039.

[21] Hans Engström, Björn Bergström, and Ulf Rosenhall. “Vestibular Sensory Epithelia”. In: Archives of Otolaryngology 100.6 (1974), pp. 411–418. DOI: 10.1001/archotol.1974.00780040425002.

[22] U. Rosenhall. “Degenerative patterns in the aging human vestibular neuro-epithelia”. In: Acta Oto-Laryngologica 76.1-6 (1973), pp. 208–220.

[23] Steven D. Rauch et al. “Decreasing Hair Cell Counts in Aging Humans”. In: Annals of the New York Academy of Sciences 942.1 (2001), pp. 220–227. DOI: 10.1111/j.1749-6632.2001.tb03748.x.

[24] Lars-Göran Johnsson and Joseph E. Hawkins. “Sensory and Neural Degeneration with Aging, as Seen in Microdissections of the Human Inner Ear”. In: Annals of Otology, Rhinology & Laryngology 81.2 (1972), pp. 179–193. DOI: 10.1177/000348947208100203.

[25] E. Richter. “Quantitative study of human Scarpa’s ganglion and vestibular sensory epithelia”. In: Acta Oto-Laryngologica 90.3-4 (1980), pp. 199–208.

[26] Robert W. Baloh and Vicente Honrubia. Clinical neurophysiology of the vestibular system. 3rd ed. New York: Oxford University Press, 2001. ISBN: 0195139828.

[27] J. G. Colebatch, S. Govender, and S. M. Rosengren. “Two distinct patterns of VEMP changes with age”. In: Clinical Neurophysiology 124.10 (May 2013), p. 2066. ISSN: 1388-2457. DOI: 10.1016/j.clinph.2013.04.337.

[28] Paul F. Smith. “The Growing Evidence for the Importance of the Otoliths in Spatial Memory”. In: Frontiers in Neural Circuits 13 (2019). DOI: 10.3389/fncir.2019.00066.

[29] Robin T. Bigelow and Yuri Agrawal. “Vestibular involvement in cognition: Visuospatial ability, attention, executive function, and memory”. In: Journal of Vestibular Research 25.2 (2015), p. 73. DOI: 10.3233/VES-150544.

[30] Kathleen E. Cullen. “The neural encoding of self-generated and externally applied movement: implications for the perception of self-motion and spatial memory”. In: Frontiers in Integrative Neuroscience 7 (2014), p. 108. DOI: 10.3389/fnint.2013.00108.

[31] Ryan M. Yoder and Jeffrey S. Taube. “The vestibular contribution to the head direction signal and navigation”. In: Frontiers in integrative neuroscience 8 (2014), p. 32. DOI: 10.3389/fnint.2014.00032. URL: https://www.ncbi.nlm.nih.gov/pubmed/24795578.

[32] Rachel E. Roditi and Benjamin T. Crane. “Suprathreshold asymmetries in human motion perception”. In: Experimental Brain Research 219.3 (2012), pp. 369–379. DOI: 10.1007/s00221-012-3099-3.

[33] E. Anson et al. “Reduced vestibular function is associated with longer, slower steps in healthy adults during normal speed walking”. In: Gait amp; posture 68 (Feb. 2019), pp. 340–345. DOI: 10.1016/j.gaitpost.2018.12.016. URL: 10.1016/j.gaitpost.2018.12.016.

[34] Katharina Hüfner et al. “GrayMatter Atrophy after Chronic Complete Unilateral Vestibular Deafferentation”. In: Annals of the New York Academy of Sciences 1164.1 (2009), p. 383. DOI: 10.1111/j.1749-6632.2008.03719.x.

[35] Peter Zu Eulenburg, Peter Stoeter, and Marianne Dieterich. “Voxelbased morphometry depicts central compensation after vestibular neuritis”. In: Annals of Neurology 68.2 (2010), p. 241. DOI: 10.1002/ana.22063.

[36] Christoph Helmchen et al. “Structural Changes in the Human Brain following Vestibular Neuritis Indicate Central Vestibular Compensation”. In: Annals of the New York Academy of Sciences 1164.1 (2009), p. 104. DOI: 10.1111/j.1749-6632.2008.03745.x.

[37] Sung-Kwang Kwang Hong et al. “Changes in the gray matter volume during compensation after vestibular neuritis: A longitudinal VBM study”. In: Restorative Neurology and Neuroscience 32.5 (2014), p. 663. DOI: 10.3233/RNN-140405.

[38] Olympia Kremmyda et al. “Beyond Dizziness: Virtual Navigation, Spatial Anxiety and Hippocampal Volume in Bilateral Vestibulopathy”. In: Frontiers in Human Neuroscience 10 (2016). DOI: 10.3389/fnhum.2016.00139.

[39] Sebastian Wurthmann et al. “Cerebral gray matter changes in persistent postural perceptual dizziness”. In: Journal of Psychosomatic Research 103 (2017), p. 95. DOI: 10.1016/j.jpsychores.2017.10.007.

[40] Martin Göttlich et al. “Hippocampal gray matter volume in bilateral vestibular failure”. In: Human Brain Mapping 37.5 (2016), p. 1998. DOI: 10.1002/hbm.23152.

[41] K. E. Hupfeld et al. “Sensory system-specific associations between brain structure and balance”. In: Neurobiology of Aging 119 (Aug. 2022), p. 102. ISSN: 0197-4580. DOI: 10.1016/j.neurobiolaging.2022.07.013.

[42] Dominic Padova et al. “Vestibular Function is Associated with Prefrontal and Sensorimotor Cortical Gray Matter Volumes in a Cross-Sectional Study of Healthy, Older Adults”. In: Aperture Neuro 4 (2024).

[43] Athira Jacob et al. “Vestibular function and cortical and sub-cortical alterations in an aging population”. In: Heliyon 6.8 (Aug. 2020), e04728. DOI: 10.1016/j.heliyon.2020.e04728. URL: 10.1016/j.heliyon.2020.e04728.

[44] Rebecca Kamil et al. “Vestibular Function and Hippocampal Volume in the Baltimore Longitudinal Study of Aging (BLSA)”. In: Otology & Neurotology 39.6 (July 2018), pp. 765–771. ISSN: 1531-7129. DOI: 10.1097/MAO.0000000000001838. URL: http://ovidsp.ovid.com/ovidweb.cgi?T=JS&NEWS=n&CSC=Y&PAGE=fulltext&D=ovft&AN=00129492-201807000-00022.

[45] Dominic M. Padova et al. “Linking vestibular function and sub-cortical grey matter volume changes in a longitudinal study of aging adults”. In: ApertureNeuro (2020). URL: https://www.humanbrainmapping.org/files/Aperture%20Neuro/Accepted%20Works%20PDF/2_39_Padovaa_Linking_vestibular_function.pdf.

[46] Nathan Wetherill Shock. Normal human aging: The Baltimore longitudinal study of aging. 84. US Department of Health and Human Services, Public Health Service, National …, 1984.

[47] S-U Ko et al. “Sex-specific age associations of ankle proprioception test performance in older adults: results from the Baltimore Longitudinal Study of Aging”. In: Age and Ageing 44.3 (2015), p. 485. DOI: 10.1093/ageing/afv005.

[48] K. D. Nguyen et al. “Test-retest reliability and age-related characteristics of the ocular and cervical vestibular evoked myogenic potential tests”. In: Otology Neurotology 31.5 (2010), p. 793. DOI: 10.1097/MAO.0b013e3181e3d60e. URL: http://search.ebscohost.com/login.aspx?direct=true&db=rzh&AN=105039616&site=ehost-live&scope=site.

[49] C. Li et al. “How to interpret latencies of cervical and ocular vestibular-evoked myogenic potentials: Our experience in fifty-three participants”. In: Clinical Otolaryngology 39.5 (2014), p. 297. DOI: 10.1111/coa.12277. URL: http://search.ebscohost.com/login.aspx?direct=true&db=asn&AN=98370966&site=ehost-live&scope=site.

[50] Carol Li et al. “Epidemiology of Vestibulo-Ocular Reflex Function”. In: Otology & Neurotology 36.2 (2015), p. 267. DOI: 10.1097/MAO.0000000000000610.

[51] Yuri Agrawal et al. “Head Impulse Test Abnormalities and Influence on Gait Speed and Falls in Older Individuals”. In: Otology amp; neurotology 34.9 (Dec. 2013), pp. 1729– 1735. DOI: 10.1097/MAO.0b013e318295313c. URL: https://www.ncbi.nlm.nih.gov/pubmed/23928523.

[52] Yuri Agrawal et al. “Evaluation of quantitative head impulse testing using search coils versus video-oculography in older individuals”. In: Otology Neurotology 35.2 (2014), p. 283. DOI: 10.1097/MAO.0b013e3182995227. URL: http://search.ebscohost.com/login.aspx?direct=true&db=rzh&AN=107881223&site=ehost-live&scope=site.

[53] Erich Schneider et al. “EyeSeeCam: An Eye Movement–Driven Head Camera for the Examination of Natural Visual Exploration”. In: Annals of the New York Academy of Sciences 1164 (2009), p. 461. DOI: 10.1111/j.1749-6632.2009.03858.x. URL: http://search.ebscohost.com/login.aspx?direct=true&db=asn&AN=40076453&site=ehost-live&scope=site.

[54] Konrad P. Weber et al. “Impulsive Testing of Semicircular-Canal Function Using Video-oculography”. In: Annals of the New York Academy of Sciences 1164 (2009), p. 486. DOI: 10.1111/j.1749-6632.2008.03730.x. URL: http://search.ebscohost.com/login.aspx?direct=true&db=asn&AN=40076497&site=ehost-live&scope=site.

[55] Laurent Younes, Marilyn Albert, and Michael I. Miller. “Inferring changepoint times of medial temporal lobe morphometric change in preclinical Alzheimer’s disease”. In: NeuroImage Clinical 5 (2014), pp. 178–187. DOI: 10.1016/j.nicl.2014.04.009.

[56] Michael I. Miller et al. “Amygdalar atrophy in symptomatic Alzheimer’s disease based on diffeomorphometry: the BIOCARD cohort”. In: Neurobiology of Aging 36 (2015), S3–S10. DOI: 10.1016/j.neurobiolaging.2014.06.032.

[57] Anqi Qiu et al. “Regional shape abnormalities in mild cognitive impairment and Alzheimer’s disease”. In: NeuroImage 45.3 (2009), pp. 656–661. DOI: 10.1016/j.neuroimage.2009.01.013.

[58] Andreia V. Faria et al. “Linking white matter and deep gray matter alterations in premanifest Huntington disease”. In: NeuroImage Clinical 11 (2016), pp. 450–460. DOI: 10.1016/j.nicl.2016.02.014.

[59] Anqi Qiu et al. “Basal Ganglia Volume and Shape in Children With Attention Deficit Hyperactivity Disorder”. In: The American Journal of Psychiatry 166.1 (2009), pp. 74–82. DOI: 10.1176/appi.ajp.2008.08030426.

[60] Anqi Qiu et al. “Hippocampal-cortical structural connectivity disruptions in schizophrenia: An integrated perspective from hippocampal shape, cortical thickness, and integrity of white matter bundles”. In: NeuroImage 52.4 (2010), pp. 1181–1189. DOI: 10.1016/j.neuroimage.2010.05.046.

[61] Anqi Qiu et al. “Region-of-interest-based analysis with application of cortical thickness variation of left planum temporale in schizophrenia and psychotic bipolar disorder”. In: Human Brain Mapping 29.8 (2008), pp. 973–985. DOI: 10.1002/hbm.20444.

[62] J Tilak Ratnanather, Chin-Fu Liu, and Michael I Miller. “Shape Diffeomorphometry of Brain Structures in Neurodegeneration and Neurodevelopment”. In: Handbook of Neuroengineering. Ed. by Nitish V. Thakor. Singapore: Springer Singapore, 2022, pp. 1–22.

[63] Dan Wu and Susumu Mori. “Structural Neuroimaging: From Macroscopic to Microscopic Scales”. In: Handbook of Neuroengineering. Ed. by Nitish V. Thakor. Singapore: Springer Singapore, 2023, pp. 2917–2951.

[64] Jun Ma, Michael I. Miller, and Laurent Younes. “A Bayesian Generative Model for Surface Template Estimation”. In: International Journal of Biomedical Imaging 2010 (2010), pp. 1–14. DOI: 10.1155/2010/974957.

[65] Denis Mongin et al. “Imputing missing data of function and disease activity in rheumatoid arthritis registers: what is the best technique?” In: RMD open 5.2 (2019), e000994.

[66] Estelle Nakul, Fabrice Bartolomei, and Christophe Lopez. “Vestibular-Evoked Cerebral Potentials”. In: Frontiers in Neurology 12 (Sept. 2021), p. 674100. ISSN: 1664-2295. DOI: 10.3389/fneur.2021.674100. URL: https://search.proquest.com/docview/2580691248.

[67] C. Lopez, O. Blanke, and F. W. Mast. “The human vestibular cortex revealed by coordinate-based activation likelihood estimation meta-analysis”. In: Neuroscience 212 (June 2012), pp. 159–179. DOI: 10.1016/j.neuroscience.2012.03.028. URL: https://www.clinicalkey.es/playcontent/1-s2.0-S0306452212002898.

[68] P. Zu Eulenburg et al. “Meta-analytical definition and functional connectivity of the human vestibular cortex”. In: NeuroImage 60.1 (2011), p. 162. DOI: 10.1016/j.neuroimage.2011.12.032.

[69] Klaus Jahn et al. “Inverse U-shaped curve for age dependency of torsional eye movement responses to galvanic vestibular stimulation”. In: Brain 126.7 (2003), p. 1579. DOI: 10.1093/brain/awg163.

[70] Peter Zu Eulenburg et al. “Ageing-related changes in the cortical processing of otolith information in humans”. In: European Journal of Neuroscience 46.12 (Nov. 2017), p. 2817. ISSN: 0953-816X. DOI: 10.1111/ejn.13755.

[71] Edmund T. Rolls et al. “Prefrontal and somatosensory-motor cortex effective connectivity in humans”. In: Cerebral Cortex 33.8 (2023), p. 4939. DOI: 10.1093/cercor/bhac391.

[72] S. Ebata et al. “Vestibular projection to the periarcuate cortex in the monkey”. In: Neuroscience Research 49.1 (2004), p. 55. DOI: 10.1016/j.neures.2004.01.012.

[73] Safiye Çavdar et al. “The brainstem connections of the supplementary motor area and its relations to the corticospinal tract: Experimental rat and human 3-tesla tractography study”. In: Neuroscience Letters 798 (2023). DOI: 10.1016/j.neulet.2023.137099.

[74] Sam J Gilbert et al. “Functional specialization within rostral prefrontal cortex (area 10): a meta-analysis”. In: Journal of Cognitive Neuroscience 18.6 (2006), pp. 932–948.

[75] Edmund T. Rolls et al. “The connectivity of the human frontal pole cortex, and a theory of its involvement in exploit versus explore”. In: Cerebral Cortex (New York, N.Y.: 1991) (Nov. 2023). DOI: 10.1093/cercor/bhad416. URL: https://search.proquest.com/docview/2892660264.

[76] Ke Peng et al. “Brodmann area 10: Collating, integrating and high level processing of nociception and pain”. In: Progress in Neurobiology 161 (Feb. 2018), pp. 1–22. ISSN: 0301-0082. DOI: 10.1016/j.pneurobio.2017.11.004. URL: https://www.ncbi.nlm.nih.gov/pubmed/29199137.

[77] Maya Danneels et al. “The impact of vestibular function on cognitive-motor interference: a case-control study on dual-tasking in persons with bilateral vestibulopathy and normal hearing”. In: Scientific Reports 13.1 (Aug. 2023), p. 13772. ISSN: 2045-2322. DOI: 10.1038/s41598-023-40465-2. URL: https://search.proquest.com/docview/2856166563.

[78] Kathleen E. Hupfeld et al. “Differential Relationships Between Brain Structure and Dual Task Walking in Young and Older Adults”. In: Frontiers in Aging Neuroscience 14 (Mar 2022). DOI: 10.3389/fnagi.2022.809281.

[79] Joyce Bosmans et al. “Associations of Bilateral Vestibulopathy With Cognition in Older Adults Matched With Healthy Controls for Hearing Status”. In: JAMA Otolaryngology– Head & Neck Surgery 148.8 (2022). DOI: 10.1001/jamaoto.2022.1303.

[80] Eric X. Wei et al. “Psychometric Tests and Spatial Navigation: Data From the Baltimore Longitudinal Study of Aging”. In: Frontiers in Neurology 11 (June 2020). DOI: 10.3389/fneur.2020.00484.

[81] Luzia Grabherr et al. “Mental transformation abilities in patients with unilateral and bilateral vestibular loss”. In: Experimental brain research 209.2 (Mar. 2011), pp. 205–214. DOI: 10.1007/s00221-011-2535-0. URL: https://link.springer.com/article/10.1007/s00221-011-2535-0.

[82] Ivan Moser et al. “Impaired math achievement in patients with acute vestibular neuritis”. In: Neuropsychologia 107 (Dec. 2017), pp. 1–8. DOI: 10.1016/j.neuropsychologia.2017.10.032. URL: 10.1016/j.neuropsychologia.2017.10.032.

[83] Nora Preuss, Fred Mast, and Gregor Hasler. “Purchase decision-making is modulated by vestibular stimulation”. In: Frontiers in behavioral neuroscience 8 (2014), p. 51.

[84] Ede A. Rancz et al. “Widespread Vestibular Activation of the Rodent Cortex”. In: The Journal of neuroscience 35.15 (Apr. 2015), pp. 5926–5934. DOI: 10.1523/JNEUROSCI.1869-14.2015. URL: https://www.ncbi.nlm.nih.gov/pubmed/25878265.

[85] Max Gattie, Elena V. M. Lieven, and Karolina Kluk. “Weak Vestibular Response in Persistent Developmental Stuttering”. In: Frontiers in Integrative Neuroscience 15 (Sept. 2021), p. 662127. ISSN: 1662-5145. DOI: 10.3389/fnint.2021.662127. URL: https://search.proquest.com/docview/2568282652.

[86] Seyede Faranak Emami et al. “Vestibular hearing and speech processing”. In: International Scholarly Research Notices 2012 (2012).

[87] Seyede Faranak Emami. “Central representation of cervical vestibular evoked myogenic potentials”. In: Indian Journal of Otolaryngology and Head & Neck Surgery 75.3 (2023), pp. 2722–2728.

[88] Lucy Stiles and Paul F. Smith. “The vestibular–basal ganglia connection: Balancing motor control”. In: Brain research 1597 (Feb. 2015), pp. 180–188. DOI: 10.1016/j.brainres.2014.11.063. URL: https://www.clinicalkey.es/playcontent/1-s2.0-S0006899314016709.

[89] Diane Deroualle and Christophe Lopez. “Toward a vestibular contribution to social cognition”. In: Frontiers in Integrative Neuroscience 8 (2014), p. 16.

[90] Eric Anson et al. “Failure on the Foam Eyes Closed Test of Standing Balance Associated With Reduced Semicircular Canal Function in Healthy Older Adults”. In: Ear Hearing 40.2 (Mar. 2020), p. 340. ISSN: 0196-0202. DOI: 10.1097/aud.0000000000000619.

[91] John Ashburner and Karl J. Friston. “Diffeomorphic registration using geodesic shooting and Gauss–Newton optimisation”. In: NeuroImage (Orlando, Fla.) 55.3 (Apr. 2011), pp. 954–967. DOI: 10.1016/j.neuroimage.2010.12.049. URL: 10.1016/j.neuroimage.2010.12.049.

[92] Christos Davatzikos. “Why voxel-based morphometric analysis should be used with great caution when characterizing group differences”. In: NeuroImage (Orlando, Fla.) 23.1 (Sept. 2004), pp. 17–20. DOI: 10.1016/j.neuroimage.2004.05.010. URL: 10.1016/j.neuroimage.2004.05.010.

[93] J. Tilak Ratnanather et al. “3D Normal Coordinate Systems for Cortical Areas”. In: Lecture Notes Series, Institute for Mathematical Sciences, National University of Singapore (2019), p. 167. DOI: 10.1142/9789811200137_0007.

[94] Casey Paquola et al. “Closing the mechanistic gap: the value of microarchitecture in understanding cognitive networks”. In: Trends in cognitive sciences 26.10 (Oct. 2022), pp. 873–886. DOI: 10.1016/j.tics.2022.07.001. URL: https://search.proquest.com/docview/2697095787.

[95] Karen E Seymour et al. “Anomalous subcortical morphology in boys, but not girls, with ADHD compared to typically developing controls and correlates with emotion dysregulation”. In: Psychiatry Research: Neuroimaging 261 (2017), pp. 20–28.

[96] Xiaoying Tang et al. “Response control correlates of anomalous basal ganglia morphology in boys, but not girls, with attention-deficit/hyperactivity disorder”. In: Behavioural Brain Research 367 (2019), pp. 117–127.

[97] William Hedley Thompson et al. “Dataset decay and the problem of sequential analyses on open datasets”. In: eLife 9 (May 2020). DOI: 10.7554/elife.53498.

[98] R. A. I. Bethlehem et al. “Brain charts for the human lifespan”. In: Nature (London) 604.7906 (Apr. 2022), pp. 525–533. DOI: 10.1038/s41586-022-04554-y. URL: https://www.narcis.nl/publication/RecordID/oai:cris.maastrichtuniversity.nl:publications%2Fc75d36bd-c842-4929-b45d-1f2001df4a1d.

[99] Daniel Zachlod et al. “Mapping Cytoarchitectonics and Receptor Architectonics to Understand Brain Function and Connectivity”. In: Biological psychiatry (1969) 93.5 (Mar. 2023), pp. 471–479. DOI: 10.1016/j.biopsych.2022.09.014. URL: 10.1016/j.biopsych.2022.09.014.

[100] Jeremy L. Smith et al. “Eagle-449: A volumetric, whole-brain compilation of brain atlases for vestibular functional MRI research”. In: Scientific Data 10.1 (2023). DOI: 10.1038/s41597-023-01938-1.

[101] Simon B. Eickhoff, B. T. Thomas Yeo, and Sarah Genon. “Imaging-based parcellations of the human brain”. In: Nature Reviews Neuroscience 19.11 (Nov. 2018), pp. 672–86.

[102] Chu-Chung Huang et al. “An extended Human Connectome Project multimodal parcellation atlas of the human cortex and subcortical areas”. In: Brain Structure and Function 227.3 (2022), pp. 763–778.

